# Thalamic white matter macrostructure and subnuclei volumes in Parkinson’s disease depression

**DOI:** 10.1101/2021.05.07.21256793

**Authors:** R Bhome, A Zarkali, JH Cole, RS Weil

## Abstract

**Objective:** Depression is a common non-motor feature of Parkinson’s disease (PD) which confers significant morbidity and is often challenging to treat. The thalamus is a key component in the basal ganglia - thalamocortical network critical to pathogenesis of PD and depression but the precise thalamic subnuclei involved in PD depression have not yet been identified and may even represent potential therapeutic targets.

**Methods:** We performed structural and diffusion weighted imaging on 76 participants with PD to evaluate the relationship between PD depression and grey and white matter thalamic subnuclear changes. We used a thalamic segmentation method to divide the thalamus into its 50 constituent subnuclei (25 each hemisphere). We used fixel based analysis of diffusion weighted imaging data to calculate mean fibre cross section (FC) for white matter tracts connected to each subnucleus and assessed volume and FC at baseline and 14-20 months follow-up. A generalised linear mixed model was used to evaluate the relationship between depression, subnuclei volume and mean FC for each of the 50 thalamic subnuclei, adjusting for age, gender, intracranial volume and time.

**Results:** We found that depression scores in PD were associated with lower right pulvinar anterior (PuA) subnucleus volume. Antidepressant use was associated with higher right PuA volume suggesting a possible protective effect of treatment. After follow-up, depression scores were associated with decreases in white matter tract macrostructure across almost all tracts connected to thalamic subnuclei.

**Conclusion:** We demonstrate that depression is associated with right thalamic PuA subnucleus volume loss and widespread thalamic white matter macrostructural changes, but that antidepressants may protect against volume loss in PD depression. Our work provides mechanistic insights for depression in PD, suggests possible benefits of actively treating depression, and a potential target for therapeutic intervention to the PuA subnucleus for PD depression.

## Introduction

Depression is a common neuropsychiatric feature of Parkinson’s Disease (PD) with a prevalence of around 35%.^1^ It is also the key health-related determinant of poor quality of life in PD.^2^ There is converging evidence to suggest that depression in PD arises primarily from the underlying neurobiology of the disorder rather than due to a psychological response to functional impairment. For example, depressive symptoms often arise in the prodromal stage prior to hallmark PD motor symptoms and genetic mutations linked to PD confer a risk of affective psychopathology.^3,4^

A clear understanding of the neural correlates of PD depression has remained elusive, with functional, structural, and nuclear imaging studies yielding broad findings.^5,6^ The frontotemporal regions, thalamus, amygdala, cerebellar white matter, hippocampus, nucleus accumbens, globus pallidus, anterior cingulate cortex and insula have all been implicated as potentially relevant brain regions and all three major monoaminergic systems are likely to be involved in the underlying pathophysiology.^5,6^

One brain structure that warrants further investigation is the thalamus. It occupies a pivotal position in the basal ganglia-thalamocortical networks that are affected both in the pathogenesis of PD and mood disorders.^7,8^ In PD depression specifically, functional magnetic resonance imaging (fMRI) studies show increased activation in the left mediodorsal thalamus,^9^ as well as increased connectivity between the left amygdala and bilateral mediodorsal thalami in depressed compared to non-depressed PD patients.^10^ Although white matter microstructural changes^11^ and noradrenergic denervation have been identified in the mediodorsal regions of the thalamus,^12^ those studies were not able to examine changes within individual thalamic subnuclei, limiting anatomical precision. The benefit of greater precision in defining neuroanatomical regions affected in PD depression is the potential of these locations as targets for therapeutic intervention, given their pivotal role in modulating brain circuits.

Previous work has demonstrated that antidepressants may modulate thalamic circuitry involving the pulvinar and mediodorsal subnuclei in depression in general.^13,14,15^ However, whether antidepressant use modifies changes caused by PD depression has not been investigated.

Until recently, analysing thalamic subnuclei morphology and the specific neural connections of individual nuclei has proven problematic as this has required manual segmentation of thalamic subnuclei which can be laborious as well as error-prone. Recently, however, a novel technique has been developed, that combines a Bayesian framework with probabilistic atlases based on ex-vivo histology and unsupervised appearance modelling to robustly segment these complex deep nuclei.^16^

Here, we investigated how levels of depression amongst PD patients influence thalamic subnucleus volume and white matter tracts connected to thalamic subnuclei. We hypothesised that depression would be associated with lower volume and altered white matter structure within thalamic subnuclei. We further examined the effects of antidepressant use on the pulvinar and mediodorsal subnuclei in PD, hypothesising that there would be volume and white matter structure changes although the expected direction was uncertain based on previous work.^14,15^

## Materials and Methods

### Participants

The study included 76 people with PD, who had been recruited to our UK centre from affiliated clinics, and 26 age-matched controls, who were spouses or recruited from a volunteer database. PD participants were recruited to the study consecutively and only excluded if they had a history of traumatic brain injury, major co-morbid psychiatric or neurological disorder, contraindication to MRI and PD duration of more than ten years. PD participants satisfied the Queen Square Brain Bank PD diagnostic criteria.^**17**^ Ethical approval was received from the Queen Square Ethics Committee (reference no. 15.LO.0476) and all participants provided written informed consent.

### Clinical Assessment

Clinical assessment was undertaken to evaluate symptoms relating to depression and anxiety as well as cognitive and disease specific measures. Depression and anxiety severity was measured using the Hospital Anxiety and Depression Scale (HADS),^**18**^ which has previously been validated for use in patients with PD.^**19**^ Cognitive testing included Mini Mental State Examination (MMSE)^20^ and Montreal Cognitive Assessment (MoCA)^21^ as measures of global cognition, as well as the Stroop^22^, digit span,^**23**^ verbal fluency,^**24**^ Graded Naming Test (GNT),^**25**^ Hooper Visual Organisation Test (HVOT),^**26**^ logical memory recall^**27**^ and Judgment of Line Orientation (JLO).^**28**^ Disease specific measures included the Movement Disorders Society Unified Parkinson’s Disease Rating Scale (MDS-UPDRS) which measures motor and non-motor aspects of disease severity,^**29**^ University of Miami Hallucinations Questionnaire (UM-PDHQ)^**30**^ evaluated hallucinations and REM Sleep Behaviour Disorder Questionnaire (RBDSQ)^**31**^ assessed sleep. Levodopa dose equivalence scores (LEDD) were calculated as described previously.^**32**^

Clinical assessments, including neuroimaging (see below), were undertaken at baseline and follow up between 14 and 20 months later (mean= 15.4 months).

### MRI data acquisition

All participants were scanned on a 3T Siemens Magnetom Prisma scanner (Siemens, Munich, Germany) with a 64-channel head coil. Diffusion weighted imaging (DWI) was acquired with the following parameters: b = 50 s/mm^2^ / 17 directions, b = 300 s/mm^2^ / 8 directions, b = 1000 s/mm^2^ / 64 directions, b = 2000 s/mm^2^ / 64 directions, 2×2×2 mm isotropic voxels, echo time: 3260 ms, repetition time: 58 ms, 72 slices, 2 mm thickness and acceleration time factor of 2. Acquisition time for DWI was approximately 10 minutes. T1-weighted data were acquired using whole head 3D magnetization prepared rapid acquisition gradient echo (MPRAGE) with the following parameters: voxel size 1mm^3^, echo time: 3.34 ms, repetition time: 2530 ms, flip angle: 7°).

### Thalamic segmentation

Image processing used the “recon-all” function in FreeSurfer v6.0.0 (http://www.freesurfer.net). This includes motion correction, normalisation of signal intensity, skull stripping, Talaraich correction, and automated segmentation of subcortical white matter and grey matter (GM) structures. Image files were then analysed in FreeSurfer v7.1.1 using the recently described thalamic segmentation technique.^**16**^ In brief, this method involves Bayesian segmentation based on a histologically derived probabilistic thalamic atlas which divides each thalamus into twenty-six subnuclei. We did not include the reticular nucleus in our analysis as it is not part of the main thalamic structure but rather forms a capsule around the lateral aspect of the thalamus separated by the external medullary lamina. This resulted in the segmentation of each thalamus into 25 constituent sub-nuclei (**see Figure 1**).

**Figure 1.**
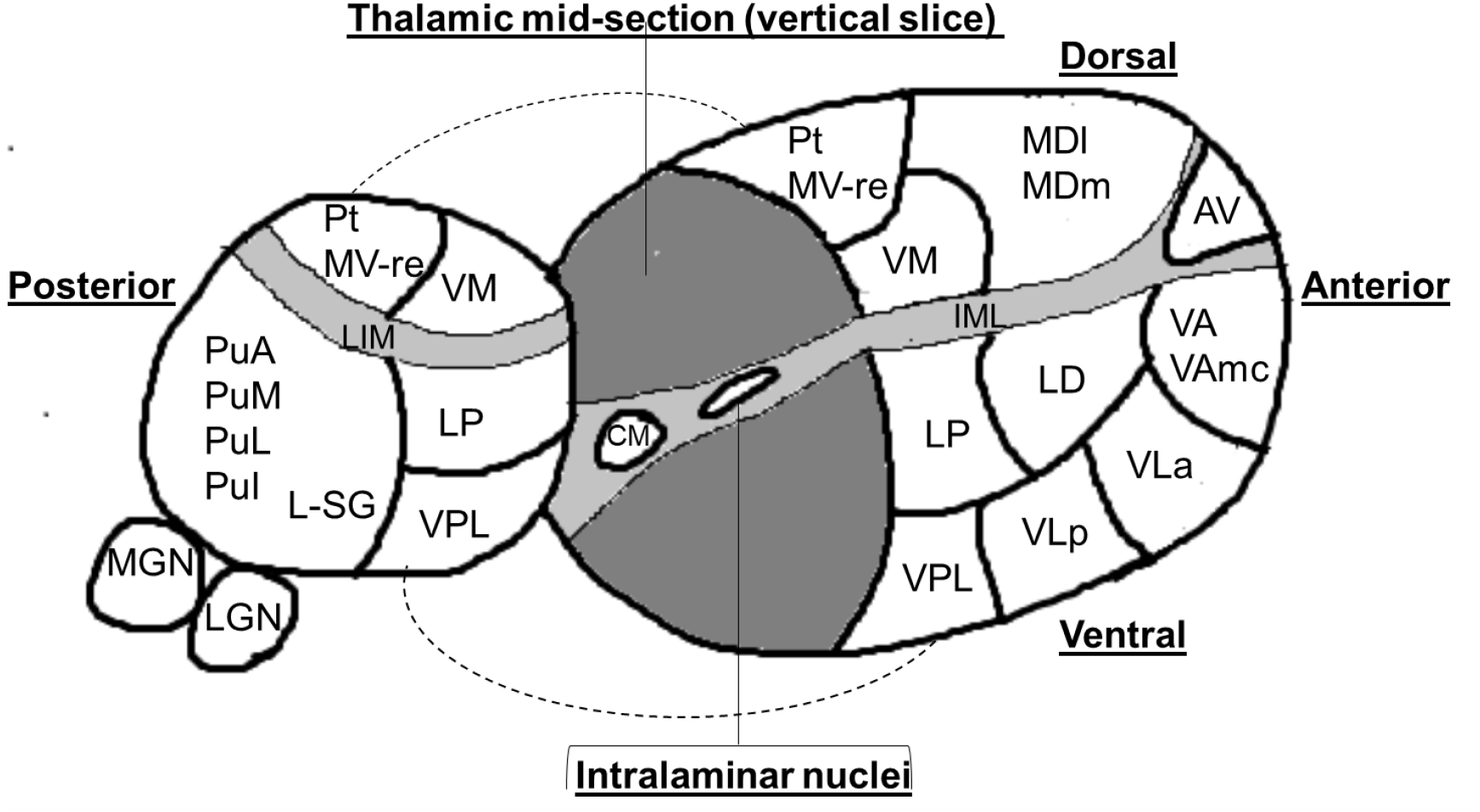
Three dimensional schematic representation of Thalamic nuclei. The thalamus is divided into two by a vertical slice to reveal the medial surface. **Abbreviations:** **Anterior nuclei** (AV= anteroventral); **Lateral nuclei (**LD= laterodorsal, LP= = lateral posterior); **Ventral nuclei** (VA= ventral anterior,VAmc= ventral anterior magnocellular, VLa= ventral lateral anterior, VLp= ventral lateral posterior, VPL= ventral posterolateral, VM= ventromedial; **Intralaminar nuclei** (CM= centromedian, CeM= central medial, CL= central lateral, Pc=paracentral, Pf= parafascicular); **Medial nuclei** (Pt= paratenial, MV-re= reuniens (medial ventral), MDm= mediodorsal medial magnocellular, MDl= mediodorsal lateral parvocellular) **Posterior nuclei** (LGN=lateral geniculate nucleus, MGN= medical geniculate nucleus, L-SG= limitans (suprageniculate), PuA= pulvinar anterior, PuM= pulvinar medial, PuL= pulvinar lateral, PuI= pulvinar inferior) IML= Internal medullary lamina

### DWI preprocessing

DWIs were denoised, corrected for Gibbs ringing, eddy-current and motion corrected and underwent bias field correction using MRtrix3 (mrtrix.org).^**33,34,35**^ In addition, DWI spatial resolution of DWIs were upsampled to a voxel size of 1.3mm^3^ and intensity normalisation was performed across participants to increase anatomic delineation and improve statistics.^**36**^ We then computed fibre-orientation distributions (FODs) for each participant using multi-shell 3-tissue constrained spherical - deconvolution, using the average response function for each tissue type (grey, white-matter, CSF).^37^ a group-averaged template was created from baseline data in 30 randomly-selected subjects (20 PD, 10 controls), and each participant’s FODs were registered to this template.

### Fibre Cross-Section

Fixel based analysis (FBA) uses a higher-order diffusion MRI model than classical diffusion tensor imaging, providing a more robust estimation of crossing fibres and allowing us to quantify degeneration within specific fibre pathways even in regions containing multiple crossing fibres.^36^ It provides information about fibre morphology as well as fibre density. Here, we focused on fibre cross section (FC) which is a relative metric of differences in fibre bundle cross section compared to a template based on the study population and is thought to be a measure of white matter macrostructure.^38^ We used this metric as our previous work in PD has shown FC to be a more sensitive measure of degeneration, compared to other fixel-based measures such as fibre density and combined fibre density and cross section.^**39**^ FC was estimated for each fixel by calculating the distortion in fibre bundle cross-section required to warp the subject image to the template image.

To assess white matter tracts connected to thalamic subnuclei, we generated tracts connected to each of the 50 thalamic subnuclei as follows: we used NiftytReg^40^ to register each subnucleus to the population template using affine linear registration. Next, we generated a tractogram for each thalamic subnucleus using probabilistic tractography on the population.^41^ We initiated streamlines within each thalamic subnucleus to the ipsilateral hemisphere, whilst excluding the rest of the thalamus (to avoid overlap between tracts). This allowed us to generate a single tract-of-interest for each thalamic subnucleus to the cortex and mean FC could then be calculated for each tract of interest in every participant.

### Statistical analysis

#### Demographics, cognitive and PD-related measures

Differences in demographics, cognitive test scores and disease-related measures were compared between participants with PD and HC. Independent sample t-tests and Mann-Whitney U tests were used for normally and non-normally distributed variables respectively, and χ^2^ for categorical variables. The Schapiro-Wilk test was used to assess normality. Statistical significance was set at p < 0.05. Statistical analyses were performed in R Version 4.0.3 and Python 3 with Jupyter Notebook version 5.5.0.

To analyse the association of depression, which was measured by the average HADS depression score at baseline and follow up, with clinical features of PD we used a General Linear Mixed Model (GLMM). The clinical measure of interest was the dependent variable and average HADS depression score, the independent variable, with time as a co-variate and participant as a random effect. For cognitive measures, age was an additional covariate while for disease specific measures, LEDD and disease duration were additional covariates. A false discovery rate (FDR) correction was performed for 19 clinical measures, using the Benjamini-Hochberg method.

#### Thalamic volumes and Fibre Cross Section

Thalamic nuclei volumes and the mean FC of fibre bundles connected to thalamic subnuclei were computed. Two separate regression models were used to evaluate the relationship between HADS-depression scores and thalamic subnuclei volume and FC for each subnucleus in PD participants. In these models subnuclei volume or mean FC were the dependent variables. For a cross-sectional evaluation at baseline, we used GLMM with baseline HADS-depression score as the independent variable, age, intracranial volume and gender as co-variates, and participant as a random effect. In a model to evaluate whether HADS-depression scores predicted follow-up nuclei volumes and FC, we used a GLMM with average HADS-depression score derived from baseline and follow-up visit scores, as the independent variable. Subnuclei volumes and mean FC values were the dependent variable in these models which also had time, age, intracranial volume and gender as co-variates, and participant as a random effect. A false discovery rate (FDR) correction was performed for 50 thalamic subnuclei tested for both volume and FC analyses, using the Benjamini-Hochberg method.

We evaluated the effect of antidepressant use on measures of volume and FC in the mediodorsal and pulvinar nuclei bilaterally by adding antidepressant use as a covariate to the GLMM outlined above.

### Data Availability

Imaging and clinical data used in this study will be shared upon reasonable request to the corresponding author. All data and statistics generated from this study are presented in the manuscript and supplementary data.

## Results

### Demographic and clinical characteristics

102 participants were included; 76 with PD and 26 HC. Age, gender and years of education did not differ between groups. At baseline, the PD group had significantly higher scores than HC on HADS anxiety and HADS depression, suggesting increased affective psychopathology (**see Table 1**), as would be expected in PD. On cognitive testing, the PD group scored significantly lower than HC on MoCA but the average score for both groups was within normal range (≥26). Scores on specific tests of language, visuospatial ability, executive functioning and memory, both immediate and delayed, did not differ between groups. The PD group had significantly higher scores than HC on disease-specific measures including total UPDRS, UPDRS motor score, and REM sleep scores, as expected but there was no significant difference in hallucinations score (**see Table 1**).

**Table 1.** Demographics, cognitive and disease specific measures of participants with PD and HC.

| Attribute | PD (n=76) | HC (n=26) | Statistic |
| --- | --- | --- | --- |
| **Demographics** |  |  |  |
| Age, y | 64.58 (7.98) | 67.42 (8.02) | t=-1.55, p=0.12 |
| Male, n (%) | 42 (55) | 12 (46) | $\chi^2=0.33$, p=0.56 |
| Education, y | 17.26 (2.85) | 17.85 (2.16) | U=884.0, p=0.42 |
| Handedness, (n, Right) (%) | 61 (80) | 24 (92) | $\chi^2=1.25$, p=0.26 |
| **Mood (HADS)** |  |  |  |
| **Depression score** | **3.95 (3.17)** | **1.15 (1.49)** | **U= 1591.5, p=2.79x10<sup>-6</sup>** |
| **Anxiety score** | **5.70 (3.77)** | **3.54 (3.43)** | **U=1333.5, p=0.008** |
| **Cognitive testing** |  |  |  |
| MMSE | 29.01 (1.18) | 29.19 (0.88) | U=953.0, p=0.78 |
| **MoCA** | **27.95 (2.20)** | **29.00 (1.14)** | **U=717.0, p=0.03** |
| Stroop time <sup>a</sup> | 62.83 (21.80) | 56.22 (14.01) | U=1144.0, p=0.19 |
| Digit Span (forwards)<sup>b</sup> | 9.25 (2.00) | 9.33 (2.05) | U=348.5, p=0.94 |
| Digit Span (backwards)<sup>b</sup> | 7.34 (2.19) | 6.92 (2.33) | t=0.59, p=0.55 |
| Log memory delayed<sup>b</sup> | 13.25 (4.48) | 12.75 (3.32) | t=0.36, p=0.72 |
| Log memory immediate<sup>b</sup> | 15.37 (4.44) | 15.17 (3.53) | t=0.15, p=0.88 |
| Word recall | 24.17 (1.10) | 24.46 (1.01) | U=810.5, p=0.13 |
| Fluency letter | 17.04 (5.17) | 17.81 (4.95) | U=910.0, p=0.55 |
| Fluency categorical | 21.72 (5.68) | 21.88 (4.66) | U=955.0, p=0.80 |
| GNT | 24.12 (2.58) | 23.62 (4.15) | U=937.5, p=0.70 |
| HVOT | 24.83 (2.96) | 25.94 (2.10) | U=769.0, p=0.09 |
| JLO<sup>a</sup> | 24.83 (3.93) | 26.0 (3.29) | t=-1.35, p=0.18 |
| **Disease specific measures** |  |  |  |
| Disease duration, y | 4.12 (2.48) |  |  |
| LEDD | 427.85 (219.66) |  |  |
| Affected side, right (%) | 36 (47) |  |  |
| left (%) | 11 (14) |  |  |
| Bilateral (%) | 29 (38) |  |  |
| **UPDRS** | **45.87 (21.01)** | **9.54 (5.71)** | **U=1954.5, p=1.17x10<sup>-13</sup>** |
| **UPDRS Motor Score** | **23.38 (12.33)** | **5.96 (4.51)** | **U=1864.5, =1.69x10<sup>-11</sup>** |
| **RBDSQ** | **4.16 (2.25)** | **1.96 (1.51)** | **U=1535.0, p=2.24X10<sup>-5</sup>** |
| UM-PDHQ | 0.68 (1.85) | 0.04 (0.19) | U=1111.0, p=0.10 |
| Abbreviations: HADS = Hospital Anxiety and Depression Scale; MMSE = Mini-Mental State Examination; MoCA = Montreal Cognitive Assessment; Stroop time = Time for completion of both word and colour tasks; Log memory = Logical memory; GNT = Graded Naming Test; HVOT = Hooper Visual Organisation Test; JLO = Judgment of Line Orientation; LEDD = Total levodopa equivalent dose; UPDRS = Unified Parkinson's Disease Rating Scale; RBDSQ = REM Sleep Behaviour Disorder Screening Questionnaire; UM-PDHQ = University of Miami Hallucinations Questionnaire<br>All data shown are mean (SD) except sex, handedness and affected side. Significant differences are highlighted in bold text.<br><sup>a</sup>PD (n=75); <sup>b</sup>PD (n=59) |  |  |  |

### Greater severity in disease measures associated with depression

In PD, at baseline, increased HADS depression score was significantly associated (post FDR correction) with higher anxiety scores (HADS) and total UPDRS scores which is an overall measure of Parkinson’s disease severity (**Table 2**). HADS depression scores did not change significantly between baseline and follow-up in PD participants (baseline, mean= 3.95 (SD=3.17); follow up, mean= 4.24 (SD=3.70), U=2829.5, p=0.83), supporting our decision to use an average HADS depression score across time points in subsequent analyses.

**Table 2.**
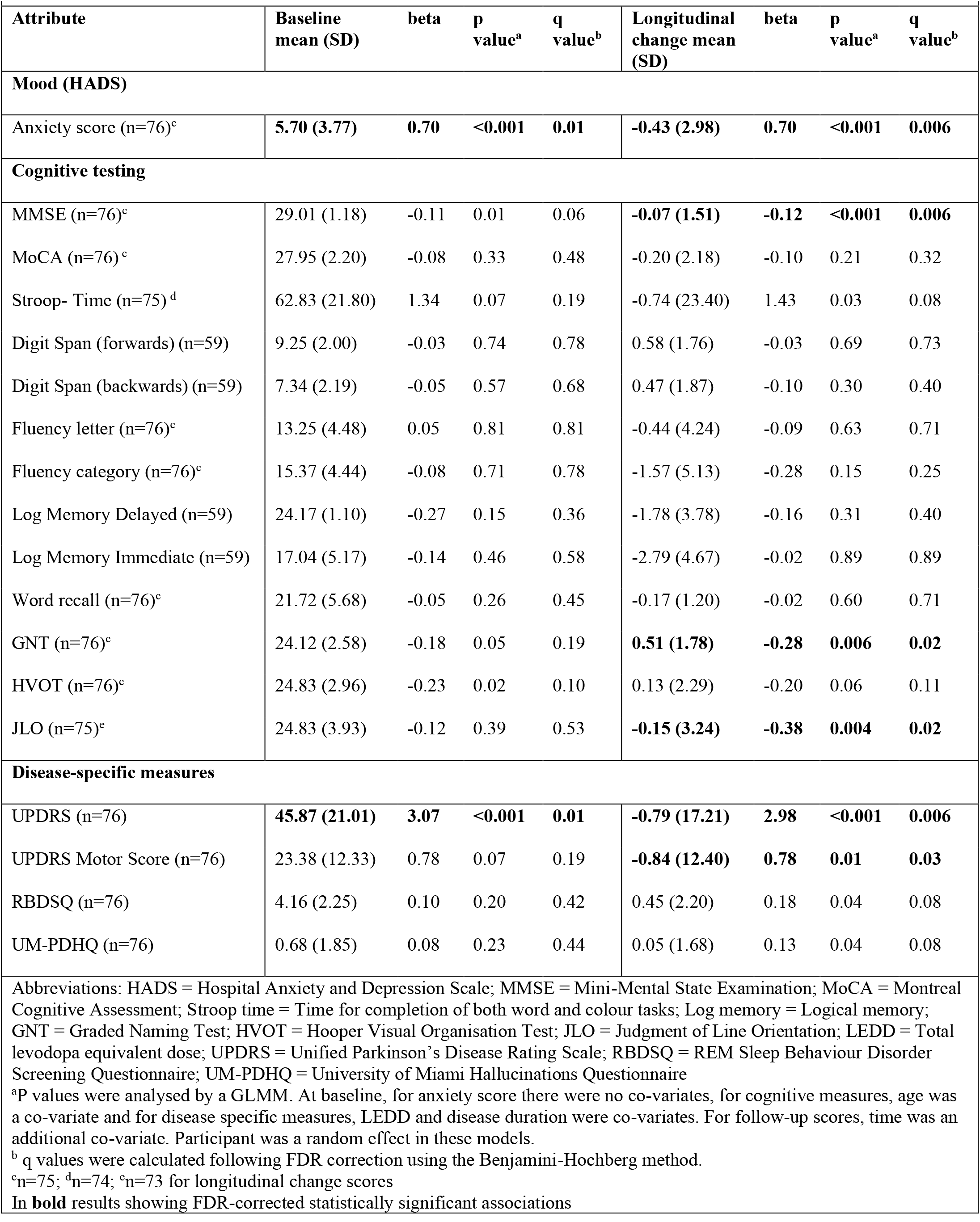
Prediction of baseline measure and follow-up scores in anxiety, cognitive and disease specific measures by HADS depression.

Average HADS depression scores predicted poorer performance at follow-up in MMSE, GNT and JLO (**Table 2**). In addition, higher depression scores also significantly predicted worse scores on the MDS-UPDRS and UPDRS motor score. However, an association between depression scores and REM sleep disorder, measured by RBDSQ, and hallucination severity, measured by UM-PDHQ, did not survive FDR correction (**Table 2**).

### Thalamic subnuclei volumes and depression

Depression severity was not significantly associated with baseline volumes of any of the thalamic subnuclei (**table e-1**). Depression scores were associated with lower right PuA volume after follow-up but this did not survive FDR correction for multiple comparisons (β=-1.32 (SE=0.52), p=0.01, q=0.50)) and with lower right PuM volume after follow-up although this was above statistical significance levels (β=-5.46 (SE=2.79), p=0.05, q=0.72) (**Table 3**).

**Table 3.**
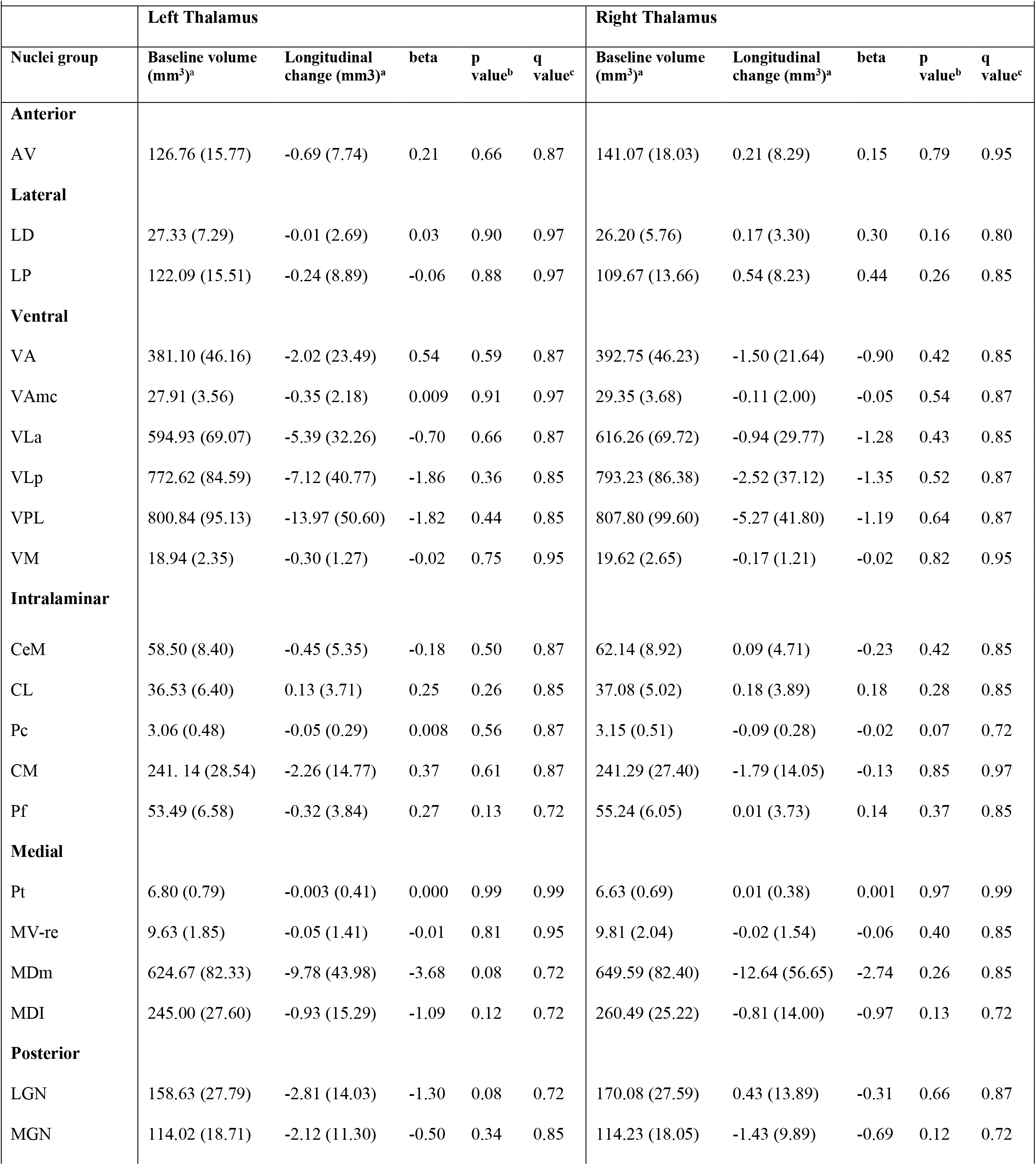
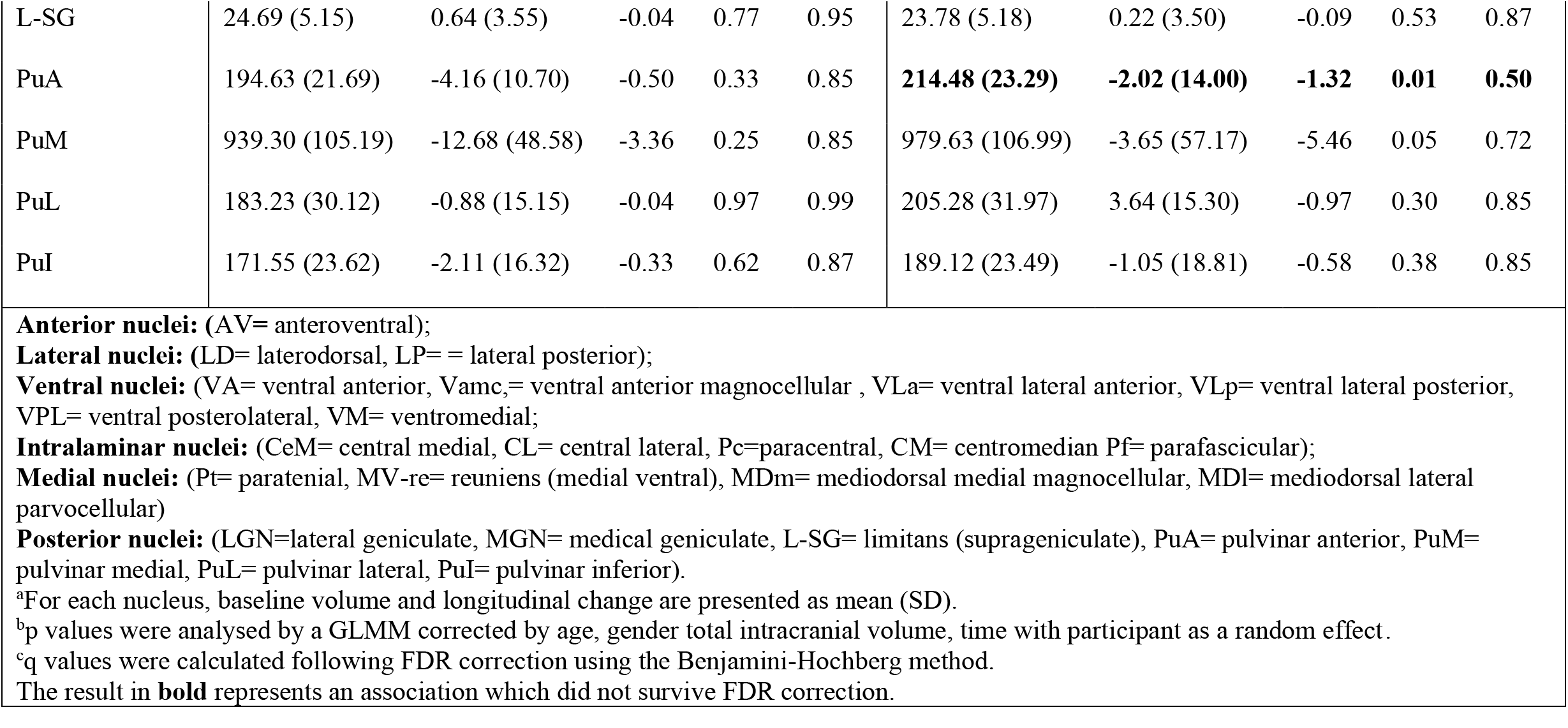
Prediction of Subnuclei volumes by HADS depression score.

In **bold** results showing FDR-corrected statistically significant associations
| Table 3. Prediction of Subnuclei volumes by HADS depression score |  |  |  |  |  |  |  |  |  |  |
| --- | --- | --- | --- | --- | --- | --- | --- | --- | --- | --- |
|  | Left Thalamus |  |  |  |  | Right Thalamus |  |  |  |  |
| Nuclei group | Baseline volume (mm<sup>3</sup>)<sup>a</sup> | Longitudinal change (mm3)<sup>a</sup> | beta | p value<sup>b</sup> | q value<sup>c</sup> | Baseline volume (mm<sup>3</sup>)<sup>a</sup> | Longitudinal change (mm<sup>3</sup>)<sup>a</sup> | beta | p value<sup>b</sup> | q value<sup>c</sup> |
| **Anterior** |  |  |  |  |  |  |  |  |  |  |
| AV | 126.76 (15.77) | -0.69 (7.74) | 0.21 | 0.66 | 0.87 | 141.07 (18.03) | 0.21 (8.29) | 0.15 | 0.79 | 0.95 |
| **Lateral** |  |  |  |  |  |  |  |  |  |  |
| LD | 27.33 (7.29) | -0.01 (2.69) | 0.03 | 0.90 | 0.97 | 26.20 (5.76) | 0.17 (3.30) | 0.30 | 0.16 | 0.80 |
| LP | 122.09 (15.51) | -0.24 (8.89) | -0.06 | 0.88 | 0.97 | 109.67 (13.66) | 0.54 (8.23) | 0.44 | 0.26 | 0.85 |
| **Ventral** |  |  |  |  |  |  |  |  |  |  |
| VA | 381.10 (46.16) | -2.02 (23.49) | 0.54 | 0.59 | 0.87 | 392.75 (46.23) | -1.50 (21.64) | -0.90 | 0.42 | 0.85 |
| VAmc | 27.91 (3.56) | -0.35 (2.18) | 0.009 | 0.91 | 0.97 | 29.35 (3.68) | -0.11 (2.00) | -0.05 | 0.54 | 0.87 |
| VLa | 594.93 (69.07) | -5.39 (32.26) | -0.70 | 0.66 | 0.87 | 616.26 (69.72) | -0.94 (29.77) | -1.28 | 0.43 | 0.85 |
| VLp | 772.62 (84.59) | -7.12 (40.77) | -1.86 | 0.36 | 0.85 | 793.23 (86.38) | -2.52 (37.12) | -1.35 | 0.52 | 0.87 |
| VPL | 800.84 (95.13) | -13.97 (50.60) | -1.82 | 0.44 | 0.85 | 807.80 (99.60) | -5.27 (41.80) | -1.19 | 0.64 | 0.87 |
| VM | 18.94 (2.35) | -0.30 (1.27) | -0.02 | 0.75 | 0.95 | 19.62 (2.65) | -0.17 (1.21) | -0.02 | 0.82 | 0.95 |
| **Intralaminar** |  |  |  |  |  |  |  |  |  |  |
| CeM | 58.50 (8.40) | -0.45 (5.35) | -0.18 | 0.50 | 0.87 | 62.14 (8.92) | 0.09 (4.71) | -0.23 | 0.42 | 0.85 |
| CL | 36.53 (6.40) | 0.13 (3.71) | 0.25 | 0.26 | 0.85 | 37.08 (5.02) | 0.18 (3.89) | 0.18 | 0.28 | 0.85 |
| Pc | 3.06 (0.48) | -0.05 (0.29) | 0.008 | 0.56 | 0.87 | 3.15 (0.51) | -0.09 (0.28) | -0.02 | 0.07 | 0.72 |
| CM | 241. 14 (28.54) | -2.26 (14.77) | 0.37 | 0.61 | 0.87 | 241.29 (27.40) | -1.79 (14.05) | -0.13 | 0.85 | 0.97 |
| Pf | 53.49 (6.58) | -0.32 (3.84) | 0.27 | 0.13 | 0.72 | 55.24 (6.05) | 0.01 (3.73) | 0.14 | 0.37 | 0.85 |
| **Medial** |  |  |  |  |  |  |  |  |  |  |
| Pt | 6.80 (0.79) | -0.003 (0.41) | 0.000 | 0.99 | 0.99 | 6.63 (0.69) | 0.01 (0.38) | 0.001 | 0.97 | 0.99 |
| MV-re | 9.63 (1.85) | -0.05 (1.41) | -0.01 | 0.81 | 0.95 | 9.81 (2.04) | -0.02 (1.54) | -0.06 | 0.40 | 0.85 |
| MDm | 624.67 (82.33) | -9.78 (43.98) | -3.68 | 0.08 | 0.72 | 649.59 (82.40) | -12.64 (56.65) | -2.74 | 0.26 | 0.85 |
| MDI | 245.00 (27.60) | -0.93 (15.29) | -1.09 | 0.12 | 0.72 | 260.49 (25.22) | -0.81 (14.00) | -0.97 | 0.13 | 0.72 |
| **Posterior** |  |  |  |  |  |  |  |  |  |  |
| LGN | 158.63 (27.79) | -2.81 (14.03) | -1.30 | 0.08 | 0.72 | 170.08 (27.59) | 0.43 (13.89) | -0.31 | 0.66 | 0.87 |
| MGN | 114.02 (18.71) | -2.12 (11.30) | -0.50 | 0.34 | 0.85 | 114.23 (18.05) | -1.43 (9.89) | -0.69 | 0.12 | 0.72 |
| L-SG | 24.69 (5.15) | 0.64 (3.55) | -0.04 | 0.77 | 0.95 | 23.78 (5.18) | 0.22 (3.50) | -0.09 | 0.53 | 0.87 |
| PuA | 194.63 (21.69) | -4.16 (10.70) | -0.50 | 0.33 | 0.85 | **214.48 (23.29)** | **-2.02 (14.00)** | **-1.32** | **0.01** | **0.50** |
| PuM | 939.30 (105.19) | -12.68 (48.58) | -3.36 | 0.25 | 0.85 | 979.63 (106.99) | -3.65 (57.17) | -5.46 | 0.05 | 0.72 |
| PuL | 183.23 (30.12) | -0.88 (15.15) | -0.04 | 0.97 | 0.99 | 205.28 (31.97) | 3.64 (15.30) | -0.97 | 0.30 | 0.85 |
| PuI | 171.55 (23.62) | -2.11 (16.32) | -0.33 | 0.62 | 0.87 | 189.12 (23.49) | -1.05 (18.81) | -0.58 | 0.38 | 0.85 |

**Anterior nuclei:** (AV= anteroventral); **Lateral nuclei:** (LD= laterodorsal, LP= = lateral posterior); **Ventral nuclei:** (VA= ventral anterior, Vamc,= ventral anterior magnocellular , VLa= ventral lateral anterior, VLp= ventral lateral posterior, VPL= ventral posterolateral, VM= ventromedial); **Intralaminar nuclei:** (CeM= central medial, CL= central lateral, Pc=paracentral, CM= centromedian Pf= parafascicular); **Medial nuclei:** (Pt= paratenial, MV-re= reuniens (medial ventral), MDm= mediodorsal medial magnocellular, MDl= mediodorsal lateral parvocellular) **Posterior nuclei:** (LGN=lateral geniculate, MGN= medical geniculate, L-SG= limitans (suprageniculate), PuA= pulvinar anterior, PuM= pulvinar medial, PuL= pulvinar lateral, PuI= pulvinar inferior). <sup>a</sup>For each nucleus, baseline volume and longitudinal change are presented as mean (SD). <sup>b</sup>p values were analysed by a GLMM corrected by age, gender total intracranial volume, time with participant as a random effect. <sup>c</sup>q values were calculated following FDR correction using the Benjamini-Hochberg method. The result in **bold** represents an association which did not survive FDR correction.

### Effects of antidepressant use on thalamic sub-nuclei

We examined the effects of antidepressant use. In our sample, 9 patients were taking antidepressants and did not differ in clinical measures from the patients who were not taking antidepressants. In particular, there were no significant differences in mean depression scores between those taking (n=9, 5.39 (SD=4.38)) and not taking antidepressants (n=67, 3.92 (SD=2.99), U=359.0, p=0.36) (**Table e-2**).

Adding antidepressant use as a co-variate in the analysis strengthened the regression model which had subnuclei volume as the dependent variable, average HADS depression score as the independent variable and time, age, intracranial volume and gender as co-variates (with antidepressant use, AIC= 1245.6; without antidepressant use, AIC= 1249.2, p=0.02). In this model, the association between depression scores and volume remained significant in the right PuA (β=-1.57 (SE=0.52), p=0.003) and became significant for the right PuM (β=-6.40 (SE=2.83), p= 0.02) at follow-up. Antidepressant use significantly predicted higher right PuA volume at follow-up (β= 12.27 (SE=5.34), p=0.02). We further found that in these subnuclei, for a higher depression score, patients taking antidepressants showed less volume loss than those not taking antidepressants but this relationship did not reach statistical significance. (**Figure 2**).

**Figure 2.**
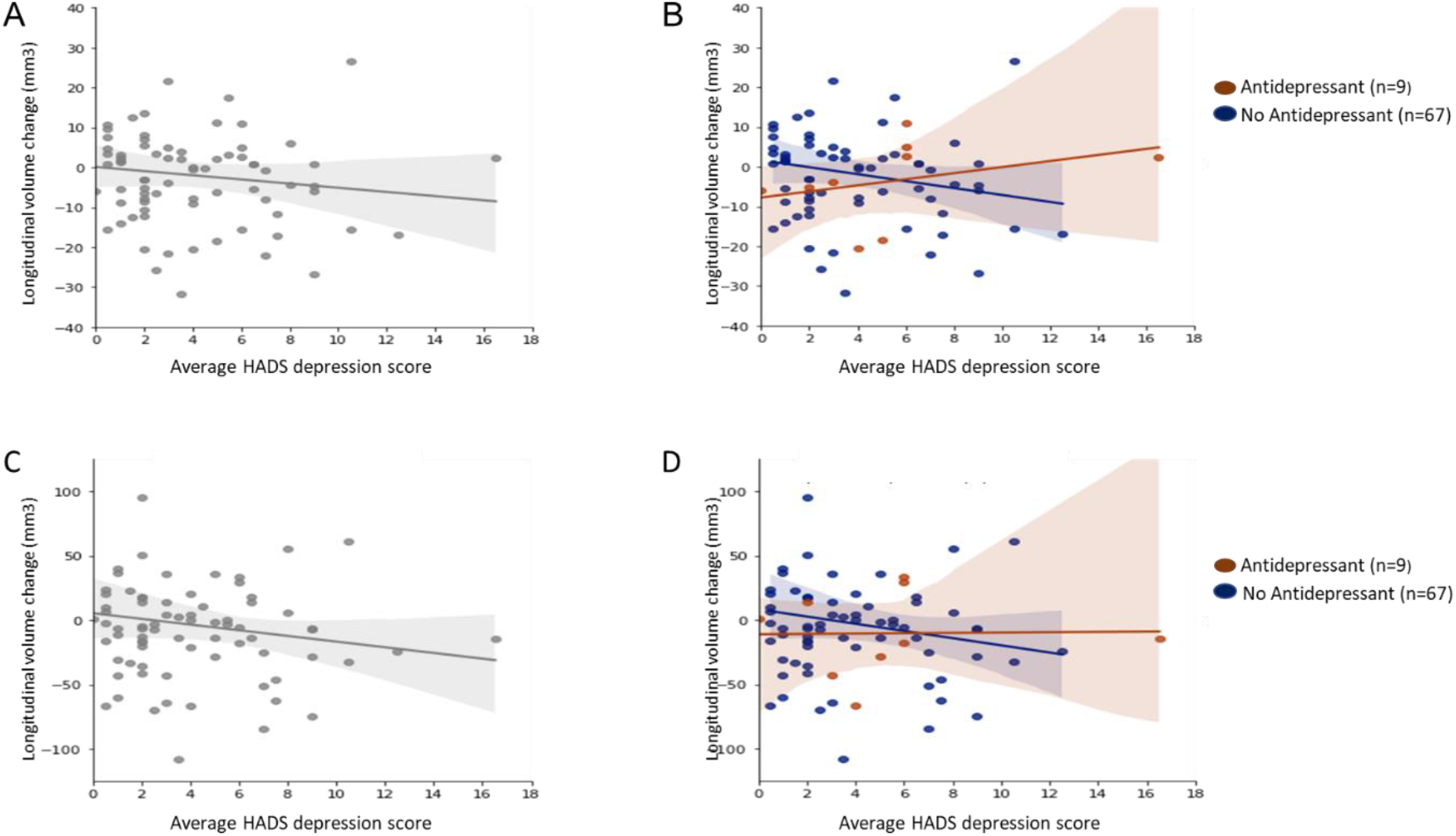
Relationship between HADS depression score and longitudinal change in pulvinar anterior and pulvinar medial volumes. The top panels show scatter plots for longitudinal change in pulvinar anterior (PuA) volume in relation to average HADS depression score for all PD participants (A) and also separately for PD participants taking (brown) and not taking (blue) antidepressants (B). The corresponding scatter plots for pulvinar medial (PuM) volume change in relation t o average HADS depression are shown in the bottom panels for all PD participants (C) and separately for participants taking (brown) or not taking (blue) antidepressants (D). Overall, higher HADS depression scores were associated with volume decreases in the right PuA (r=-0.12, P=0.30) (A) and right PuM (r=-0.12, P=0.28) (C). However, when participants were separated into those taking or not taking antidepressants, there was a positive correlation between HADS depression scores and longitudinal volume change in the right PuA for those taking antidepressants (r=0.34, P=0.37) but a negative correlation for those not taking antidepressants (r=-0.18, p=0.14) (B). There was a similar pattern for the right PuM; Antidepressant group (r=0.02, P=0.96); No antidepressant group (r=-0.14, P=0.26) (D).

### Fibre cross section changes associated with depression

At baseline, there were no significant relationships between depression scores and FC (**Table e-3**). After follow-up, we found a significant association between HADS depression scores and FC for tracts connected to all thalamic subnuclei apart from the left PuA, left Pt and right Pt (**Table 4; Table e-4**). This relationship was of reduced FC with higher depression scores for all tracts apart from the VM and Pc nuclei bilaterally, which showed the opposite relationship, with increased FC with higher depression severity. All significant associations survived correction for multiple comparisons.

**Table 4.** Prediction of follow-up FC scores by average HADS depression score.

|  |  |  |  |  |  |  |  |  |  |  |
| --- | --- | --- | --- | --- | --- | --- | --- | --- | --- | --- |
| Left Thalamus |  |  |  |  |  | Right Thalamus |  |  |  |  |
| Nuclei group | Baseline FC<sup>a</sup> | Longitudinal change<sup>a</sup> | beta | p value<sup>b</sup> | q value<sup>c</sup> | Baseline FC<sup>a</sup> | Longitudinal change<sup>a</sup> | beta | p value<sup>b</sup> | q value<sup>c</sup> |
| **Anterior** |  |  |  |  |  |  |  |  |  |  |
| AV | 0.071 | -0.197 | -0.011 | 0.018 | 0.026 | 0.078 | -0.199 | -0.012 | 0.014 | 0.026 |
| **Lateral** |  |  |  |  |  |  |  |  |  |  |
| LD | 0.085 | -0.198 | -0.012 | 0.015 | 0.026 | 0.083 | -0.199 | -0.012 | 0.013 | 0.026 |
| LP | 0.016 | -0.168 | -0.012 | 0.007 | 0.026 | 0.075 | -0.180 | -0.012 | 0.010 | 0.026 |
| **Ventral** |  |  |  |  |  |  |  |  |  |  |
| VA | 0.070 | -0.202 | -0.011 | 0.026 | 0.029 | 0.071 | -0.205 | -0.011 | 0.017 | 0.026 |
| VAmc | 0.067 | -0.196 | -0.011 | 0.018 | 0.026 | 0.072 | -0.206 | -0.011 | 0.017 | 0.026 |
| VLa | 0.067 | -0.202 | -0.011 | 0.028 | 0.026 | 0.066 | -0.206 | -0.011 | 0.020 | 0.026 |
| VLp | 0.066 | -0.202 | -0.010 | 0.030 | 0.033 | 0.067 | -0.205 | -0.011 | 0.021 | 0.026 |
| VPL | 0.069 | -0.200 | -0.011 | 0.025 | 0.028 | 0.065 | -0.202 | -0.011 | 0.021 | 0.026 |
| VM | -0.300 | 0.833 | 0.043 | 0.031 | 0.033 | -0.287 | 0.834 | 0.044 | 0.020 | 0.026 |
| **Intralaminar** |  |  |  |  |  |  |  |  |  |  |
| CeM | 0.065 | -0.194 | -0.011 | 0.020 | 0.026 | 0.068 | -0.197 | -0.011 | 0.019 | 0.026 |
| CL | 0.083 | -0.194 | -0.011 | 0.014 | 0.026 | 0.085 | -0.205 | -0.014 | 0.017 | 0.026 |
| Pc | -0.217 | 0.585 | 0.034 | 0.017 | 0.026 | -0.227 | 0.605 | 0.036 | 0.014 | 0.026 |
| CM | 0.070 | -0.198 | -0.011 | 0.019 | 0.026 | 0.073 | -0.202 | -0.011 | 0.017 | 0.026 |
| Pf | 0.062 | -0.190 | -0.011 | 0.018 | 0.026 | 0.069 | -0.196 | -0.011 | 0.018 | 0.026 |
| **Medial** |  |  |  |  |  |  |  |  |  |  |
| Pt | -0.217 | -0.217 | 0.009 | 0.080 | 0.083 | -0.227 | -0.227 | 0.009 | 0.120 | 0.12 |
| MV-re | 0.069 | -0.196 | -0.011 | 0.017 | 0.026 | 0.074 | -0.202 | -0.011 | 0.016 | 0.026 |
| MDm | 0.039 | -0.168 | -0.011 | 0.010 | 0.026 | 0.039 | -0.177 | -0.010 | 0.020 | 0.026 |
| MDI | 0.016 | -0.161 | -0.011 | 0.012 | 0.026 | -0.007 | -0.110 | -0.010 | 0.007 | 0.026 |
| **Posterior** |  |  |  |  |  |  |  |  |  |  |
| LGN | 0.081 | -0.207 | -0.011 | 0.022 | 0.026 | 0.074 | -0.199 | -0.012 | 0.013 | 0.026 |
| MGN | 0.080 | -0.200 | -0.011 | 0.018 | 0.026 | 0.077 | -0.199 | -0.012 | 0.015 | 0.026 |
| L-SG | 0.069 | -0.183 | -0.011 | 0.014 | 0.026 | 0.090 | -0.195 | -0.012 | 0.010 | 0.026 |
| PuA | -0.016 | -0.160 | -0.010 | 0.097 | 0.099 | **0.047** | **-0.169** | **-0.012** | **0.007** | **0.026** |
| PuM | **0.084** | **-0.201** | **-0.011** | **0.017** | **0.026** | **0.080** | **-0.198** | **-0.012** | **0.013** | **0.026** |
| PuL | **0.076** | **-0.204** | **-0.011** | **0.025** | **0.028** | **0.072** | **-0.205** | **-0.011** | **0.018** | **0.026** |
| PuI | **0.083** | **-0.205** | **-0.012** | **0.017** | **0.026** | **0.081** | **-0.204** | **-0.012** | **0.014** | **0.026** |

**Abbreviations:**
**Anterior nuclei:** (AV= anteroventral);
**Lateral nuclei:** (LD= laterodorsal, LP= = lateral posterior);
**Ventral nuclei:** (VA= ventral anterior, Vamc,= ventral anterior magnocellular , VLa= ventral lateral anterior, VLp= ventral lateral posterior, VPL= ventral posterolateral, VM= ventromedial;
**Intralaminar nuclei:** (CeM= central medial, CL= central lateral, Pc=paracentral, CM= centromedian Pf= parafascicular);
**Medial nuclei:** (Pt= paratenial, MV-re= reuniens (medial ventral), MDm= mediodorsal medial magnocellular, MDl= mediodorsal lateral parvocellular)
**Posterior nuclei:** (LGN=lateral geniculate, MGN= medical geniculate, L-SG= limitans (suprageniculate), PuA= pulvinar anterior, PuM= pulvinar medial, PuL= pulvinar lateral, PuI= pulvinar inferior).
<sup>a</sup> For each nucleus, baseline FC and longitudinal change are presented as mean (SD).
<sup>b</sup>p values were analysed by a GLMM corrected by age, gender total intracranial volume, time with participant as a random effect.
<sup>c</sup>q values were calculated following FDR correction using the Benjamini-Hochberg method.

## Discussion

We examined the effects of Parkinson’s depression on thalamic subnuclei employing a histologically - based automated MRI segmentation method. This allowed us to analyse individual thalamic subnuclei volumes and white matter connections of these subnuclei. We found that lower volume in the right pulvinar, specifically the right PuA subnucleus, is predicted by depression scores and that antidepressant use was related to higher right PuA volume. Additionally, we found that depression scores were associated with reduced FC across almost all thalamic subnuclei at follow-up. This suggests that depression in PD is associated with widespread loss of white matter macrostructural integrity of fibres projecting to and from the thalamus. Notably, we also found that depression in Parkinson’s disease was associated with worse total UPDRS scores, motor symptoms, anxiety, MMSE scores, visuospatial performance and naming ability.

A detailed analysis of structural changes in thalamic subnuclei in PD has not been previously reported, but right-sided pulvinar dysfunction has been implicated in depressive illness, more generally, outside of the context of PD.^14,15^ Interestingly, treatment of major depressive disorder with duloxetine, a potentially effective antidepressant in PD depression,^42^ strengthens functional connectivity between the right pulvinar and right orbitofrontal cortex and also between the right pulvinar and the limbic regions of the right anterior cingulate cortex, left dorsomedial prefrontal cortex and left temporal gyrus.^15^ These regions, along with the default mode network, to which the right pulvinar is connected regulate affective control and are implicated in mood disorders.^**43,44**,^ Intriguingly, we found that antidepressant use was associated with higher right PuA and PuM volumes at follow-up compared to participants not taking antidepressants. Given that PD depression is likely to arise from disruption and degeneration of neural networks implicated in non-PD depression,^5^ our findings, in conjunction with the broader literature, implicate the right PuA as a subnucleus of interest in the aetiology of PD depression.

Severity of PD depression was associated with reduced FC across the majority of thalamic nuclei suggesting widespread white matter tract atrophy amongst fibres connected to the thalamus. Previous work has highlighted depression as a marker of PD disease severity.^**45**^ Therefore, the association between depression scores and widespread white matter atrophy of tracts involving thalamic nuclei may be indirect. Indeed, alpha-synuclein deposition and neuronal loss have been consistently demonstrated in thalamic nuclei in PD and are likely to be correlated with disease severity.^46^

In contrast, we saw increased FC after follow-up associated with higher depression scores in the bilateral VM and Pc nuclei. The VM nuclei have uniquely strong and diffuse interconnections to widespread cortical regions including the prefrontal cortex, enabling them to dynamically synchronise cortical networks.^**47**^ It is plausible that white matter hypertrophy in the VM nuclei represents a compensatory mechanism to offset dysregulated neural circuits in brain regions implicated in PD depression, for example, the pulvinar-cortical networks. Similarly, white matter hypertrophy of tracts originating from the Pc may represent neural plasticity in response to aberrant neural circuity in affective control pathways involving the prefrontal cortex. However, these subnuclei had lower probabilistic values than other subnuclei, and it is possible that with smaller subnuclei with lower value estimations, tract estimations are less robust, making any conclusions linked to higher FC in these subnuclei tentative. Further work will be needed to validate these findings in PD depression in other cohorts.

Depression scores significantly predicted poorer clinical features of PD including motor symptoms even after correcting for dopamine dose and disease duration. Similarly, depression scores also predicted poorer MMSE, naming ability and visuospatial performance, after correcting for age. The relationship between depressive symptomatology and severity of motor symptoms is in keeping with several previous studies.^,45,48^ The exact mechanism by which depression influences motor functioning remains unclear although a recent study of de novo PD patients found that depressed patients already had more severe motor symptoms at baseline, suggesting that depression is a marker of disease severity. Interestingly, in that study, both depressed and non-depressed PD patients had similar striatal dopaminergic levels suggesting that in PD depression, factors other than dopamine levels could affect motor symptoms.^**49**^ We found that depression was also associated with visuoperceptual deficits which have also been shown to be associated with widespread macrostructural white matter degeneration.^**39**^ It is most likely that PD depression is linked with these other non-motor PD features due to a shared aetiology of accelerated and more widespread Lewy body deposition in brain regions beyond the midbrain dopaminergic neurons.

There are some potential limitations to consider. Depression was measured using a single measure, the HADS. Using a combination of multiple tools to measure depression severity could strengthen validity and reliability. We examined relationships with depression severity scores, rather than separating depressed from non-depressed patients, due to limited availability of clinical data on depression. Our cohort did not include patients with more severe depressive symptoms. Future work should examine thalamic changes in PD across the spectrum of depression severity, to determine whether these findings apply across this range, or mainly relate to milder depressive symptoms.

Participants with other pathologies such as hypertension that could influence white matter structure and potentially affect FC values were not excluded; and similar to previous studies using fixel based analysis, white matter hyperintensities were not specifically quantified or controlled for.^**50**^

## Conclusion

Our study utilised a novel thalamic segmentation tool to investigate volume and white matter macrostructural changes associated with depressive symptomatology in PD. Novel findings include PD depression being associated with right PuA volume loss and widespread thalamic white matter macrostructural loss as well as antidepressant use being associated with higher right PuA volume. Future work should examine PD patients with more severe depression and over a longer follow-up. In the longer term, a better understanding of the neural correlates of PD depression may reveal potential therapeutic targets.

## Acknowledgements

The authors thank all the participants for their time.

### Appendix

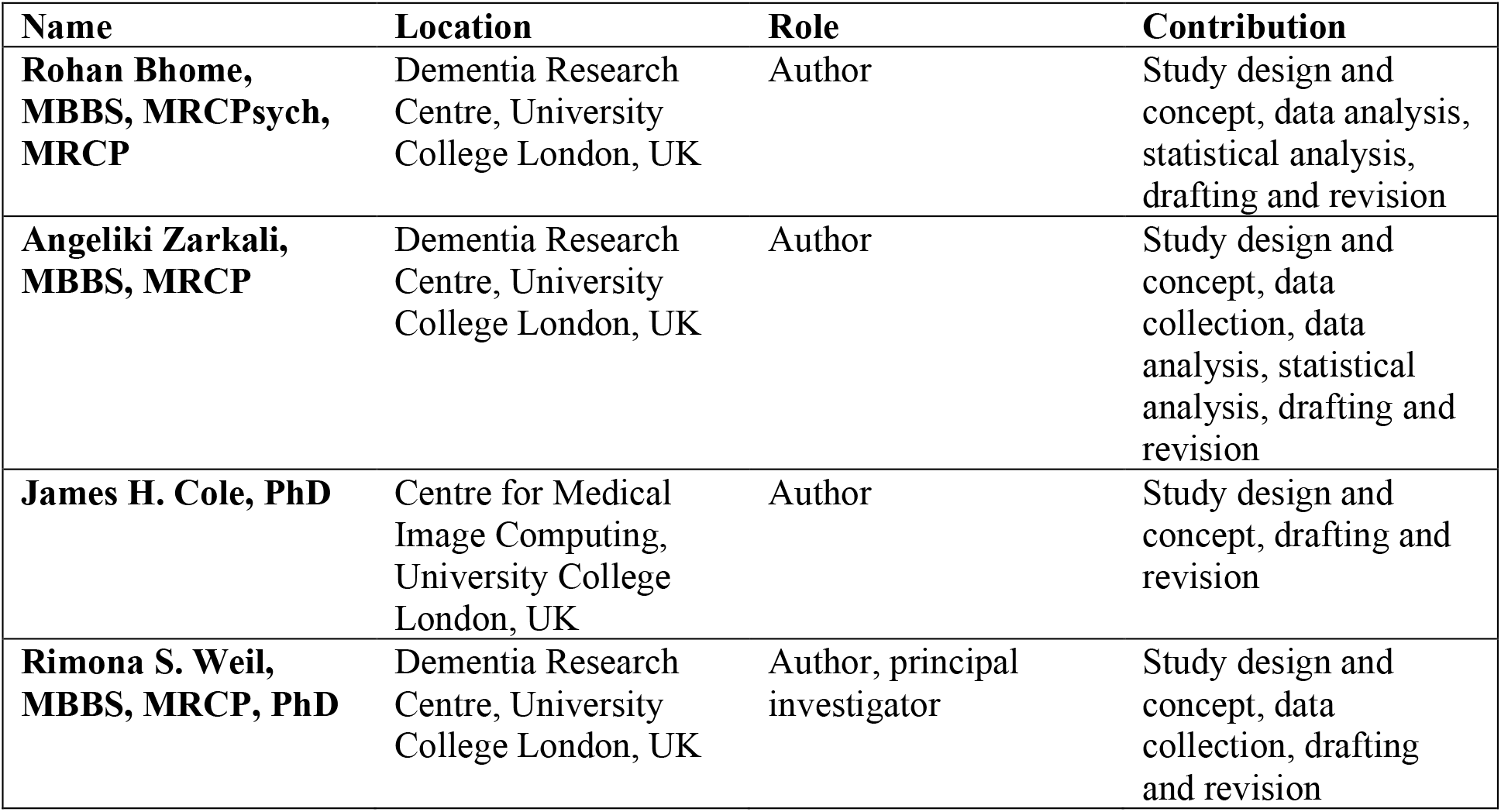

| **Name** | **Location** | **Role** | **Contribution** |
| --- | --- | --- | --- |
| **Rohan Bhome, MBBS, MRCPsych, MRCP** | Dementia Research Centre, University College London, UK | Author | Study design and concept, data analysis, statistical analysis, drafting and revision |
| **Angeliki Zarkali, MBBS, MRCP** | Dementia Research Centre, University College London, UK | Author | Study design and concept, data collection, data analysis, statistical analysis, drafting and revision |
| **James H. Cole, PhD** | Centre for Medical Image Computing, University College London, UK | Author | Study design and concept, drafting and revision |
| **Rimona S. Weil, MBBS, MRCP, PhD** | Dementia Research Centre, University College London, UK | Author, principal investigator | Study design and concept, data collection, drafting and revision |

