## Supplementary material for "Thalamic white matter macrostructure and subnuclei volumes in Parkinson’s disease depression": Table e-1

### Title

| Supplementary Table 1. Association between baseline thalamic nuclei volumes and HADS depression score | | | | | | | | |
|---|---|---|---|---|---|---|---|---|
| | **Left Thalamus** | | | | **Right Thalamus** | | | |
| **Nuclei** | **Baseline Volume (mm$^3$)** | **Adjusted beta (SE)** | **95% CI** | **p value[a]** | **Baseline Volume (mm$^3$)** | **Adjusted beta (SE)** | **95% CI** | **p value[a]** |
| **Anterior group** | | | | | | | | |
| AV | 126.76 (15.77) | 0.11 (0.49) | -0.86 – 1.07 | 0.83 | 141.07 (18.03) | -0.17 (0.61) | -1.36 – 1.03 | 0.79 |
| **Lateral group** | | | | | | | | |
| LD | 27.33 (7.29) | 0.05 (0.27) | -0.49 – 0.58 | 0.87 | 26.20 (5.76) | 0.30 (0.21) | -0.11 – 0.71 | 0.16 |
| LP | 122.09 (15.51) | -0.27 (0.48) | -1.21 – 0.66 | 0.57 | 109.67 (13.66) | 0.58 (0.42) | -0.25 – 1.41 | 0.17 |
| **Ventral group** | | | | | | | | |
| VA | 381.10 (46.16) | -0.43 (1.13) | -2.66 – 1.79 | 0.70 | 392.75 (46.23) | -2.07 (1.21) | -4.43 – 0.29 | 0.09 |
| VAmc | 27.91 (3.56) | -0.09 (0.10) | -0.28 – 0.10 | 0.35 | 29.35 (3.68) | -0.12 (0.10) | -0.32 – 0.07 | 0.22 |
| VLa | 594.93 (69.07) | -1.71 (1.69) | -5.02 – 1.60 | 0.31 | 616.26 (69.72) | -2.99 (1.80) | -6.52 – 0.54 | 0.10 |
| VLp | 772.62 (84.59) | -2.91 (2.25) | -7.33 – 1.50 | 0.20 | 793.23 (86.38) | -3.07 (2.27) | -7.52 – 1.37 | 0.18 |
| VPL | 800.84 (95.13) | -3.44 (2.65) | -8.63 – 1.75 | 0.19 | 807.80 (99.60) | -1.77 (2.74) | -7.13 – 3.59 | 0.52 |
| VM | 18.94 (2.35) | -0.05 (0.07) | -0.18 – 0.09 | 0.51 | 19.62 (2.65) | -0.04 (0.08) | -0.18 – 0.11 | 0.64 |
| **Intralaminar group** | | | | | | | | |
| CeM | 58.50 (8.40) | -0.16 (0.28) | -0.71 – 0.40 | 0.58 | 62.14 (8.92) | -0.33 (0.30) | -0.92 – 0.26 | 0.27 |
| CL | 36.53 (6.40) | 0.28 (0.23) | -0.17 – 0.73 | 0.22 | 37.08 (5.02) | 0.19 (0.18) | -0.16 – 0.54 | 0.29 |
| Pc | 3.06 (0.48) | -0.01 (0.01) | -0.04 – 0.02 | 0.42 | 3.15 (0.51) | -0.03 (0.02) | -0.06 – 0.004 | 0.10 |
| CM | 241. 14 (28.54) | -0.04 (0.02) | -0.05 – 0.04 | 0.87 | 241.29 (27.40) | -0.53 (0.76) | -2.00 – 0.96 | 0.49 |
| Pf | 53.49 (6.58) | 0.15 (0.19) | -0.23 – 0.52 | 0.44 | 55.24 (6.05) | 0.15 (0.17) | -0.19 – 0.49 | 0.39 |
| **Medial group** | | | | | | | | |
| Pt | 6.80 (0.79) | -0.004 (0.02) | -0.05 – 0.04 | 0.87 | 6.63 (0.69) | -0.001 (0.02) | -0.04 – 0.04 | 0.94 |
| MV-re | 9.63 (1.85) | -0.02 (0.06) | -0.15 – 0.10 | 0.74 | 9.81 (2.04) | -0.06 (0.07) | -0.20 – 0.08 | 0.40 |
| MDm | 624.67 (82.33) | -4.63 (2.42) | -9.38 – 0.12 | 0.06 | 649.59 (82.40) | -3.21 (2.42) | -7.95 – 1.54 | 0.19 |
| MDl | 245.00 (27.60) | -1.46 (0.80) | -3.02 – 0.11 | 0.07 | 260.49 (25.22) | -0.82 (0.71) | -2.20 – 0.56 | 0.25 |
| **Posterior group** | | | | | | | | |
| LGN | 158.63 (27.79) | -1.32 (0.80) | -2.88 – 0.24 | 0.10 | 170.08 (27.59) | -0.20 (0.80) | -1.77 – 1.37 | 0.80 |
| MGN | 114.02 (18.71) | -0.69 (0.60) | -1.87 – 0.49 | 0.25 | 114.23 (18.05) | -0.82 (0.52) | -1.83 – 0.20 | 0.12 |
| L-SG | 24.69 (5.15) | -0.05 (0.17) | -0.38 – 0.28 | 0.77 | 23.78 (5.18) | 0.05 (0.16) | -0.27 – 0.36 | 0.78 |
| PuA | 194.63 (21.69) | -0.29 (0.57) | -1.41 – 0.83 | 0.61 | 214.48 (23.29) | -0.77 (0.62) | -1.99 – 0.45 | 0.21 |

| | | | | | | | | |
|---|---|---|---|---|---|---|---|---|
| PuM | 939.30 (105.19) | -3.05 (3.10) | -9.13 – 3.03 | 0.33 | 979.63 (106.99) | -3.28 (3.19) | -9.53 – 2.97 | 0.30 |
| PuL | 183.23 (30.12) | -0.16 (0.95) | -2.02 – 1.71 | 0.87 | 205.28 (31.97) | -0.79 (1.00) | -2.75 – 1.17 | 0.43 |
| PuI | 171.55 (23.62) | -0.61 (0.69) | -1.97 – 0.76 | 0.38 | 189.12 (23.49) | -0.27 (0.73) | -1.69 – 1.16 | 0.72 |

**Anterior nuclei: (AV=** anteroventral);
**Lateral nuclei: (**LD= laterodorsal, LP= = lateral posterior);
**Ventral nuclei:** (VA= ventral anterior, Vamc,= ventral anterior magnocellular , VLa= ventral lateral anterior, VLp= ventral lateral posterior, VPL= ventral posterolateral, VM= ventromedial;
**Intralaminar nuclei:** (CeM= central medial, CL= central lateral, Pc=paracentral, CM= centromedian Pf= parafascicular);
**Medial nuclei:** (Pt= paratenial, MV-re= reuniens (medial ventral), MDm= mediodorsal medial magnocellular, MDl= mediodorsal lateral parvocellular)
**Posterior nuclei:** (LGN=lateral geniculate, MGN= medical geniculate, L-SG= limitans (suprageniculate), PuA= pulvinar anterior, PuM= pulvinar medial, PuL= pulvinar lateral, PuI= pulvinar inferior).
[a]P values were analysed by a general linear model corrected by age, gender, total intracranial volume at baseline
There were no significant associations at baseline

| **Supplementary Table 2. Characteristics of PD participants taking and not taking antidepressant medication** | | | |
|---|---|---|---|
| **Attribute** | **Antidepressant (n=9)** | **No antidepressant (n=67)** | **Statistic** |
| **Demographics** | | | |
| Age, y | 60.56 (6.47) | 65.12 (8.01) | t=-1.62, p=0.11 |
| Male, n (%) | 3 (33%) | 67 (88) | χ2=1.11, p=0.29 |
| **Mood (HADS)[a]** | | | |
| Depression score | 5.39 (4.38) | 3.92 (2.99) | U=359.0, p=0.36 |
| Anxiety score | 8.94 (5.90) | 5.01 (2.96) | U= 469.5, p=0.007 |
| **Cognitive testing** | | | |
| MMSE | 29.00 (1.15) | 29.01 (1.18) | U=299.0, p=0.97 |
| MoCA | 28.11 (1.91) | 27.93 (2.23) | U=314.5, p=0.84 |
| GNT | 24.67 (2.94) | 24.04 (2.52) | U=356.0, p=0.38 |
| JLO[b] | 24.22 (3.74) | 24.90 (3.95) | t=-0.49, p=0.63 |
| **Disease specific measures** | | | |
| Disease duration, y | 3.56 (2.01) | 4.19 (2.53) | U=260.5, p=0.51 |
| LEDD | 420.56 (178.43) | 428.83 (224.61) | t=-0.10, p=0.92 |
| UPDRS | 46.89 (13.94) | 45.73 (21.78) | U=329.5, p=0.66 |
| UPDRS Motor Score | 25.67 (10.55) | 23.07 (12.52) | U=352.5, p=0.42 |
| RBDSQ | 5.33 (3.02) | 4.00 (2.07) | U=370.5, p=0.27 |
| UM-PDHQ | 2.22 (3.29) | 0.48 (1.44) | U=372.0, p=0.08 |

Abbreviations: HADS = Hospital Anxiety and Depression Scale; MMSE = Mini-Mental State Examination; MoCA = Montreal Cognitive Assessment; GNT = Graded Naming Test; = Judgment of Line Orientation; LEDD = Total levodopa equivalent dose; UPDRS = Unified Parkinson's Disease Rating Scale; RBDSQ = REM Sleep Behaviour Disorder Screening Questionnaire; UM-PDHQ = University of Miami Hallucinations Questionnaire

All data shown are mean (SD) except gender.

[a]Average HADS depression and anxiety scores derived from baseline and follow up scores

[b] No antidepressant (n=66)

| Supplementary Table 3. Association between baseline thalamic nuclei FC and HADS depression score | | | | | | | | |
|---|---|---|---|---|---|---|---|---|
| | **Left Thalamus** | | | | **Right Thalamus** | | | |
| **Nuclei** | **Baseline FC** | **Adjusted beta (SE)** | **95% CI** | **p value[a]** | **Baseline FC** | **Adjusted beta (SE)** | **95% CI** | **p value[a]** |
| **Anterior group** | | | | | | | | |
| AV | 0.071 (0.025) | -0.002 (0.001) | -0.004 – 0.001 | 0.21 | 0.078 (0.064) | -0.002 (0.001) | -0.004 – 0.001 | 0.18 |
| **Lateral group** | | | | | | | | |
| LD | 0.085 (0.065) | -0.002 (0.001) | -0.004 – 0.001 | 0.16 | 0.083 (0.065) | -0.002 (0.001) | -0.004 – 0.001 | 0.16 |
| LP | 0.016 (0.054) | -0.000 (0.001) | -0.003 – 0.003 | 0.79 | 0.075 (0.063) | -0.002 (0.001) | -0.004 – 0.001 | 0.14 |
| **Ventral group** | | | | | | | | |
| VA | 0.070 (0.065) | -0.001 (0.001) | -0.004 – 0.001 | 0.35 | 0.071 (0.066) | -0.001 (0.001) | -0.004 – 0.001 | 0.29 |
| VAmc | 0.067 (0.059) | -0.001 (0.001) | -0.003 – 0.001 | 0.47 | 0.072 (0.063) | -0.001 (0.001) | -0.004 – 0.001 | 0.34 |
| VLa | 0.067 (0.061) | -0.001 (0.001) | -0.003 – 0.002 | 0.56 | 0.066 (0.062) | -0.001 (0.001) | -0.003 – 0.002 | 0.51 |
| VLp | 0.066 (0.061) | -0.001 (0.001) | -0.003 – 0.002 | 0.56 | 0.067 (0.061) | -0.001 (0.001) | -0.003 – 0.002 | 0.63 |
| VPL | 0.069 (0.059) | -0.001 (0.001) | -0.003 – 0.002 | 0.60 | 0.065 (0.058) | -0.000 (0.001) | -0.002 – 0.002 | 0.83 |
| VM | -0.300 (0.247) | 0.002 (0.003) | -0.008 – 0.012 | 0.66 | -0.287 (0.250) | 0.003 (0.005) | -0.007 – 0.012 | 0.55 |
| **Intralaminar group** | | | | | | | | |
| CeM | 0.065 (0.058) | -0.001 (0.001) | -0.003 – 0.002 | 0.56 | 0.068 (0.057) | -0.000 (0.001) | -0.002 – 0.002 | 0.92 |
| CL | 0.083 (0.065) | -0.002 (0.001) | -0.005 – 0.000 | 0.09 | 0.085 (0.074) | -0.003 (0.003) | -0.009 – 0.002 | 0.24 |
| Pc | -0.217 (0.180) | 0.004 (0.003) | -0.003 – 0.010 | 0.29 | -0.227 (0.144) | 0.004 (0.004) | -0003 – 0.011 | 0.27 |
| CM | 0.070 (0.060) | -0.001 (0.001) | -0.003 – 0.001 | 0.39 | 0.073 (0.060) | -0.001 (0.001) | -0.003 – 0.002 | 0.53 |
| Pf | 0.062 (0.057) | -0.001 (0.001) | -0.003 – 0.002 | 0.62 | 0.069 (0.057) | -0.000 (0.001) | -0.003 – 0.002 | 0.83 |
| **Medial group** | | | | | | | | |
| Pt | -0.217 (0.180) | 0.004 (0.003) | -0.003 – 0.001 | 0.29 | -0.227 (0.144) | 0.004 (0.004) | -0.003 – 0.011 | 0.27 |
| MV-re | 0.069 (0.059) | -0.001 (0.001) | -0.003 – 0.001 | 0.42 | 0.074 (0.062) | -0.001 (0.001) | -0.004 – 0.001 | 0.30 |
| MDm | 0.039 (0.065) | -0.002 (0.001) | -0005 – 0.001 | 0.12 | 0.039 (0.057) | 0.000 (0.002) | -0.003 – 0.003 | 0.92 |
| MDI | 0.016 (0.059) | -0.001 (0.001) | -0.004 – 0.002 | 0.50 | -0.007 (0.073) | -0.001 (0.001) | -0.005 – 0.004 | 0.78 |
| **Posterior group** | | | | | | | | |
| LGN | 0.081 (0.066) | -0.001 (0.001) | -0.004 – 0.001 | 0.25 | 0.074 (0.065) | -0.002 (0.001) | -0.004 – 0.001 | 0.16 |
| MGN | 0.080 (0.063) | -0.001 (0.001) | -0.004 – 0.001 | 0.28 | 0.077 (0.063) | -0.001 (0.001) | -0.004 – 0.001 | 0.24 |
| L-SG | 0.069 (0.065) | -0.002 (0.001) | -0.005 – 0.000 | 0.09 | 0.090 (0.066) | -0.002 (0.001) | -0.005 – 0.000 | 0.07 |
| PuA | -0.016 (0.202) | -0.004 (0.005) | -0.014 – 0.007 | 0.50 | 0.047 (0.071) | -0.002 (0.002) | -0.006 – 0.001 | 0.15 |
| PuM | 0.084 (0.066) | -0.002 (0.001) | -0.004 – 0.001 | 0.16 | 0.080 (0.066) | -0.002 (0.001) | -0.004 – 0.001 | 0.14 |

| | | | | | | | | |
|---|---|---|---|---|---|---|---|---|
| PuL | 0.076 (0.064) | -0.001 (0.001) | -0.003 – 0.001 | 0.39 | 0.072 (0.064) | -0.001 (0.001) | -0.004 – 0.001 | 0.35 |
| PuI | 0.083 (0.064) | -0.001 (0.001) | -0.004 – 0.001 | 0.25 | 0.081 (0.064) | -0.001 (0.001) | -0.004 – 0.001 | 0.23 |

**Anterior nuclei: (AV=** anteroventral);
**Lateral nuclei: (**LD= laterodorsal, LP= = lateral posterior);
**Ventral nuclei:** (VA= ventral anterior, Vamc,= ventral anterior magnocellular , VLa= ventral lateral anterior, VLp= ventral lateral posterior, VPL= ventral posterolateral, VM= ventromedial;
**Intralaminar nuclei:** (CeM= central medial, CL= central lateral, Pc=paracentral, CM= centromedian Pf= parafascicular);
**Medial nuclei:** (Pt= paratenial, MV-re= reuniens (medial ventral), MDm= mediodorsal medial magnocellular, MDl= mediodorsal lateral parvocellular)
**Posterior nuclei:** (LGN=lateral geniculate, MGN= medical geniculate, L-SG= limitans (suprageniculate), PuA= pulvinar anterior, PuM= pulvinar medial, PuL= pulvinar lateral, PuI= pulvinar inferior).
[a]P values were analysed by a general linear model corrected by age, gender, total intracranial volume at baseline
There were no significant associations at baseline

| Supplementary Table 4. Prediction of FC scores by average HADS depression score | | | | | | | | | | |
|---|---|---|---|---|---|---|---|---|---|---|
| | Left Thalamus | | | | | Right Thalamus | | | | |
| **Nuclei** | **Baseline FC[a]** | **Longitudinal change[a]** | **Adjusted beta (SE)** | **p value[b]** | **q value[c]** | **Baseline FC[a]** | **Longitudinal change[a]** | **Adjusted beta (SE)** | **p value[b]** | **q value[c]** |
| **Anterior group** | | | | | | | | | | |
| AV | **0.071 (0.025)** | **-0.197 (0.253)** | **-0.011 (0.005)** | **0.018** | **0.026** | **0.078 (0.064)** | **-0.199 (0.257)** | **-0.012 (0.005)** | **0.014** | **0.026** |
| **Lateral group** | | | | | | | | | | |
| LD | **0.085 (0.065)** | **-0.198 (0.261)** | **-0.012 (0.005)** | **0.015** | **0.026** | **0.083 (0.065)** | **-0.199 (0.262)** | **-0.012 (0.005)** | **0.013** | **0.026** |
| LP | **0.016 (0.054)** | **-0.168 (0.236)** | **-0.012 (0.004)** | **0.007** | **0.026** | **0.075 (0.063)** | **-0.180 (0.257)** | **-0.012 (0.005)** | **0.010** | **0.026** |
| **Ventral group** | | | | | | | | | | |
| VA | **0.070 (0.065)** | **-0.202 (0.258)** | **-0.011 (0.005)** | **0.026** | **0.029** | **0.071 (0.066)** | **-0.205 (0.259)** | **-0.011 (0.005)** | **0.017** | **0.026** |
| VAmc | **0.067 (0.059)** | **-0.196 (0.259)** | **-0.011 (0.005)** | **0.018** | **0.026** | **0.072 (0.063)** | **-0.206 (0.260)** | **-0.011 (0.005)** | **0.017** | **0.026** |
| VLa | **0.067 (0.061)** | **-0.202 (0.259)** | **-0.011 (0.005)** | **0.028** | **0.026** | **0.066 (0.062)** | **-0.206 (0.261)** | **-0.011 (0.005)** | **0.020** | **0.026** |
| VLp | **0.066 (0.061)** | **-0.202 (0.257)** | **-0.010 (0.005)** | **0.030** | **0.033** | **0.067 (0.061)** | **-0.205 (0.262)** | **-0.011 (0.005)** | **0.021** | **0.026** |
| VPL | **0.069 (0.059)** | **-0.200 (0.261)** | **-0.011 (0.005)** | **0.025** | **0.028** | **0.065 (0.058)** | **-0.202 (0.261)** | **-0.011 (0.005)** | **0.021** | **0.026** |
| VM | **-0.300 (0.247)** | **0.833 (1.064)** | **0.043 (0.020)** | **0.031** | **0.033** | **-0.287 (0.250)** | **0.834 (1.057)** | **0.044 (0.019)** | **0.020** | **0.026** |
| **Intralaminar group** | | | | | | | | | | |
| CeM | **0.065 (0.058)** | **-0.194 (0.258)** | **-0.011 (0.005)** | **0.020** | **0.026** | **0.068 (0.057)** | **-0.197 (0.260)** | **-0.011 (0.005)** | **0.019** | **0.026** |
| CL | **0.083 (0.065)** | **-0.194 (0.256)** | **-0.011 (0.005)** | **0.014** | **0.026** | **0.085 (0.074)** | **-0.205 (0.282)** | **-0.014 (0.006)** | **0.017** | **0.026** |
| Pc | **-0.217 (0.180)** | **0.585 (0.772)** | **0.034 (0.014)** | **0.017** | **0.026** | **-0.227 (0.144)** | **0.605 (0.803)** | **0.036 (0.015)** | **0.014** | **0.026** |
| CM | **0.070 (0.060)** | **-0.198 (0.260)** | **-0.011 (0.005)** | **0.019** | **0.026** | **0.073 (0.060)** | **-0.202 (0.262)** | **-0.011 (0.005)** | **0.017** | **0.026** |
| Pf | **0.062 (0.057)** | **-0.190 (0.256)** | **-0.011 (0.005)** | **0.018** | **0.026** | **0.069 (0.057)** | **-0.196 (0.260)** | **-0.011 (0.005)** | **0.018** | **0.026** |
| **Medial group** | | | | | | | | | | |
| Pt | -0.217 (0.180) | -0.217 (0.180) | 0.009 (0.005) | 0.080 | 0.083 | -0.227 (0.144) | -0.227 (0.144) | 0.009 (0.006) | 0.120 | 0.12 |
| MV-re | **0.069 (0.059)** | **-0.196 (0.260)** | **-0.011 (0.005)** | **0.017** | **0.026** | **0.074 (0.062)** | **-0.202 (0.260)** | **-0.011 (0.005)** | **0.016** | **0.026** |
| MDm | **0.039 (0.065)** | **-0.168 (0.230)** | **-0.011 (0.004)** | **0.010** | **0.026** | **0.039 (0.057)** | **-0.177 (0.239)** | **-0.010 (0.005)** | **0.020** | **0.026** |
| MDI | **0.016 (0.059)** | **-0.161 (0.215)** | **-0.011 (0.004)** | **0.012** | **0.026** | **-0.007 (0.073)** | **-0.110 (0.202)** | **-0.010 (0.004)** | **0.007** | **0.026** |
| **Posterior group** | | | | | | | | | | |
| LGN | **0.081 (0.066)** | **-0.207 (0.261)** | **-0.011 (0.005)** | **0.022** | **0.026** | **0.074 (0.065)** | **-0.199 (0.255)** | **-0.012 (0.005)** | **0.013** | **0.026** |
| MGN | **0.080 (0.063)** | **-0.200 (0.262)** | **-0.011 (0.005)** | **0.018** | **0.026** | **0.077 (0.063)** | **-0.199 (0.259)** | **-0.012 (0.005)** | **0.015** | **0.026** |
| L-SG | **0.069 (0.065)** | **-0.183 (0.248)** | **-0.011 (0.005)** | **0.014** | **0.026** | **0.090 (0.066)** | **-0.195 (0.265)** | **-0.012 (0.005)** | **0.010** | **0.026** |
| PuA | -0.016 (0.202) | -0.160 (0.263) | -0.010 (0.006) | 0.097 | 0.099 | **0.047 (0.071)** | **-0.169 (0.256)** | **-0.012 (0.004)** | **0.007** | **0.026** |
| PuM | **0.084 (0.066)** | **-0.201 (0.261)** | **-0.011 (0.005)** | **0.017** | **0.026** | **0.080 (0.066)** | **-0.198 (0.260)** | **-0.012 (0.005)** | **0.013** | **0.026** |

| | | | | | | | | | | |
|---|---|---|---|---|---|---|---|---|---|---|
| PuL | **0.076 (0.064)** | **-0.204 (0.261)** | **-0.011 (0.005)** | **0.025** | **0.028** | **0.072 (0.064)** | **-0.205 (0.261)** | **-0.011 (0.005)** | **0.018** | **0.026** |
| PuI | **0.083 (0.064)** | **-0.205 (0.265)** | **-0.012 (0.005)** | **0.017** | **0.026** | **0.081 (0.064)** | **-0.204 (0.263)** | **-0.012 (0.005)** | **0.014** | **0.026** |

**Anterior nuclei: (**AV= anteroventral);
**Lateral nuclei: (**LD= laterodorsal, LP= = lateral posterior);
**Ventral nuclei:** (VA= ventral anterior, Vamc,= ventral anterior magnocellular , VLa= ventral lateral anterior, VLp= ventral lateral posterior, VPL= ventral posterolateral, VM= ventromedial;
**Intralaminar nuclei:** (CeM= central medial, CL= central lateral, Pc=paracentral, CM= centromedian Pf= parafascicular);
**Medial nuclei:** (Pt= paratenial, MV-re= reuniens (medial ventral), MDm= mediodorsal medial magnocellular, MDl= mediodorsal lateral parvocellular)
**Posterior nuclei:** (LGN=lateral geniculate, MGN= medical geniculate, L-SG= limitans (suprageniculate), PuA= pulvinar anterior, PuM= pulvinar medial, PuL= pulvinar lateral, PuI= pulvinar inferior).
[a] For each nucleus, baseline FC and longitudinal change are presented as mean (SD).
[b]p values were analysed by a GLMM corrected by age, gender total intracranial volume, time with participant as a random effect.
[c]q values were calculated following FDR correction using the Benjamini-Hochberg method.
In **bold** results showing FDR-corrected statistically significant associations
